# Vaccination with BNT162b2 reduces transmission of SARS-CoV-2 to household contacts in Israel

**DOI:** 10.1101/2021.07.13.21260393

**Authors:** Ottavia Prunas, Joshua L. Warren, Forrest W. Crawford, Sivan Gazit, Tal Patalon, Daniel M. Weinberger, Virginia E. Pitzer

**Author notes:** These authors contributed equally to this work (second authors) (Joshua L. Warren, Forrest W. Crawford). These authors contributed equally to this work (senior authors) (Daniel M. Weinberger, Virginia E. Pitzer).

## Abstract

The individual-level effectiveness of vaccines against clinical disease caused by SARS-CoV-2 is well-established. However, few studies have directly examined the effect of COVID-19 vaccines on transmission. We quantified the effectiveness of vaccination with BNT162b2 (Pfizer-BioNTech mRNA-based vaccine) against household transmission of SARS-CoV-2 in Israel. We fit two time-to-event models – a mechanistic transmission model and a regression model – to estimate vaccine effectiveness against susceptibility to infection and infectiousness given infection in household settings. Vaccine effectiveness against susceptibility to infection was 80-88%. For breakthrough infections among vaccinated individuals, the vaccine effectiveness against infectiousness was 41-79%. The overall vaccine effectiveness against transmission was 88.5%. Vaccination provides substantial protection against susceptibility to infection and slightly lower protection against infectiousness given infection, thereby reducing transmission of SARS-CoV-2 to household contacts.

**One-Sentence Summary:** Vaccination reduced both the rate of infection with SARS-CoV-2 and transmission to household contacts in Israel.

## Main Text

The COVID-19 pandemic caused by SARS-CoV-2 has led to unprecedented disruptions worldwide. The rapid development and deployment of vaccines against the virus has provided an opportunity to control the outbreak in populations with access to vaccination. Multiple vaccines against SARS-CoV-2 have been demonstrated to be effective in preventing clinical disease and reducing disease severity in those who do become infected [1-4]. This direct protection against disease is critical. However, additional population-level benefits can be derived if vaccines also reduce transmission of the virus, thereby providing protection to those who are still vulnerable to infection [1, 5].

To date, there is little direct real-world evidence about the effects of vaccination on SARS-CoV-2 transmission. A few studies have investigated the reduction in transmission in households and amongst healthcare workers [4, 6]. Other studies have indirectly found evidence for a likely effect of the vaccine on transmission by demonstrating reduced viral load in the upper respiratory tract of infected individuals [7-11].

Households are an ideal setting for evaluating transmission of the virus and the effects of vaccination due to the high rate of secondary infection among household members [4, 12]. Detailed data on household structure and timing of infections can be used to quantify the risk of transmission. We aimed to assess the effectiveness of vaccination against susceptibility to infection and against infectiousness given infection with SARS-CoV-2 following vaccination with BNT162b2 (Pfizer-BioNTech mRNA-based vaccine). We accomplished this using two different analytic approaches applied to data from the second-largest healthcare organization in Israel. The rapid and early rollout of mass vaccination in Israel provides a unique opportunity to evaluate the effectiveness of the vaccine against transmission.

We used data from Maccabi Healthcare Services (MHS) centralized database, which captures all data on members’ demographics and healthcare-related interactions. MHS is a nationwide 2.5 million-member state-mandated, not-for-profit sick fund in Israel, representing a quarter of the Israeli population, and is a representative sample of the Israeli population. The full dataset, covering the period from June 15, 2020 to March 24, 2021, included information on 2,305,704 individuals from 1,275,015 households. Among these, 1,276,311 individuals received two doses of BNT162b2 as of March 24, 2021. There were 191,138 detected infections caused by SARS-CoV-2 (8.3% of the total population), with 4,141 infections following the second dose of the vaccine and 73,582 infections in unvaccinated individuals (naïve risk ratio = 5.6%).

Most of the households (60.7% of the total) had a single household member; this individual was infected in 59,552 (7.7%) of the 774,003 households. Information on the number of households and proportion of infections occurring in households of varying size can be found in table S1. We focused our analysis on households with at least one infected individual and two or more household members, for a total of 65,624 households and 253,564 individuals (see supplementary materials, materials and methods).

To infer transmission rates, it is necessary to estimate when each individual within a household was infected and the period when they were infectious. We therefore used a data augmentation approach to impute when a person with a positive PCR test was infected and infectious. This was accomplished using random samples from three different Gamma distributions representing the delay between onset of infectiousness and the date of the PCR test, the date of infection and the onset of infectiousness (i.e., latent period), and the onset of infectiousness to the end of infectiousness (i.e., infectious period) (Fig. 1 and table S2; supplementary materials, materials and methods).

**Fig. 1.**
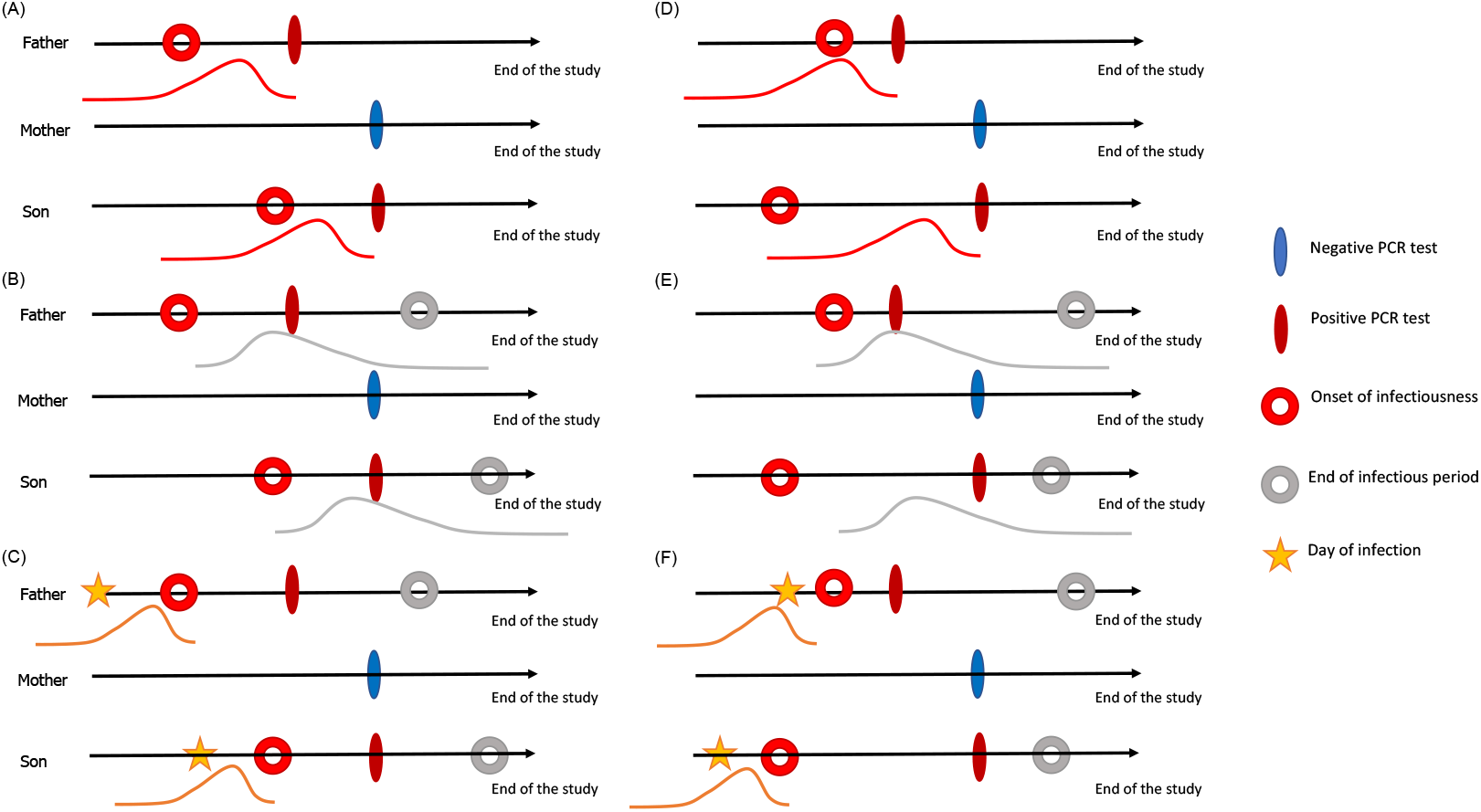
Schematic representation of the data augmentation process for an example household. Each infected household member is associated with: (**A**,**D**) a distribution for time from onset of infectiousness to testing; (**B**,**E**) a distribution for the infectious period; and (**C**,**F**) a distribution for the latent period. The filled ovals represent observed events, while the circles and stars represent unobserved events in the infection timeline. Panels (A-C) and (D-F) represent two possible sample sets from the delay distributions, each with a different index case.

We developed two discrete time-to-event data models of household transmission to estimate vaccine effectiveness against susceptibility to infection and against infectiousness given infection. In both approaches, we model the infection status for person *j* in household *i* on study date *t* (*Y*_*ijt*_) using conditionally independent Bernoulli distributions with corresponding probability of infection *π*_*ijt*_. These probabilities are then defined based on personal demographics, community risk, vaccination status, and characteristics of household transmission, with the approaches differing in how transmission is described. Both models were fit 100 times with the different draws from the delay distributions to assess uncertainty in the results due to uncertainty in the infection timeline.

Using the model of household transmission, we estimated that receipt of two doses of the vaccine was associated with an age-adjusted vaccine effectiveness against susceptibility to infection (*VE*_*S*_) of 80.5% (95% confidence interval (CI): 78.9%, 82.1%) and a vaccine effectiveness against infectiousness given infection (*VE*_*I*_) of 41.3% (95% CI: 9.5%, 73.0%). The vaccine effectiveness against transmission (*VE*_*T*_), which combines the reduction in the risk of infection and the risk of infectiousness given infection among vaccinated individuals, was estimated to be 88.5% (95% CI: 82.3%, 94.8%). Vaccine effectiveness estimates across age groups are shown in Table S3 and coefficients from the primary model are shown in Table S4.

Using the alternative infection-hazard approach, in the absence of infected household members, vaccination was associated with a reduction in the hazard of infection of *VE*_*S*,0_ = 87.9% (95% CI: 86.7%, 89.0%). If exposed to an infected, unvaccinated household member, vaccination was associated with a *VE*_*S,u*_ = 92.3% (95% CI: 90.2%, 94.5%) reduction in the hazard of infection, whereas if exposed to an infected, fully vaccinated household member, vaccination reduced the individual hazard of infection by *VE*_*S,v*_ = 64.9% (95% CI: 35.2%, 94.5%).

Amongst unvaccinated individuals, there was a *VE*_*I,u*_ = 78.6% (95% CI: 74.5%, 82.7%) reduction in the hazard of infection when exposed to a fully vaccinated versus unvaccinated infected household contact. However, the vaccination status of infected household contacts was not significantly associated with the hazard of infection amongst fully vaccinated individuals (*VE*_*I,v*_ = 3.23%; 95% CI: -0.87%, 15.7%).

We observed limited variability in the vaccine effectiveness estimates across 100 iterations of the delay distributions (Fig. 2 and 3), suggesting that our results are robust to the unobserved time-course of infection within individuals. Furthermore, we found robust results when including both vaccine doses in the two models (table S5 and S6).

**Fig. 2.**
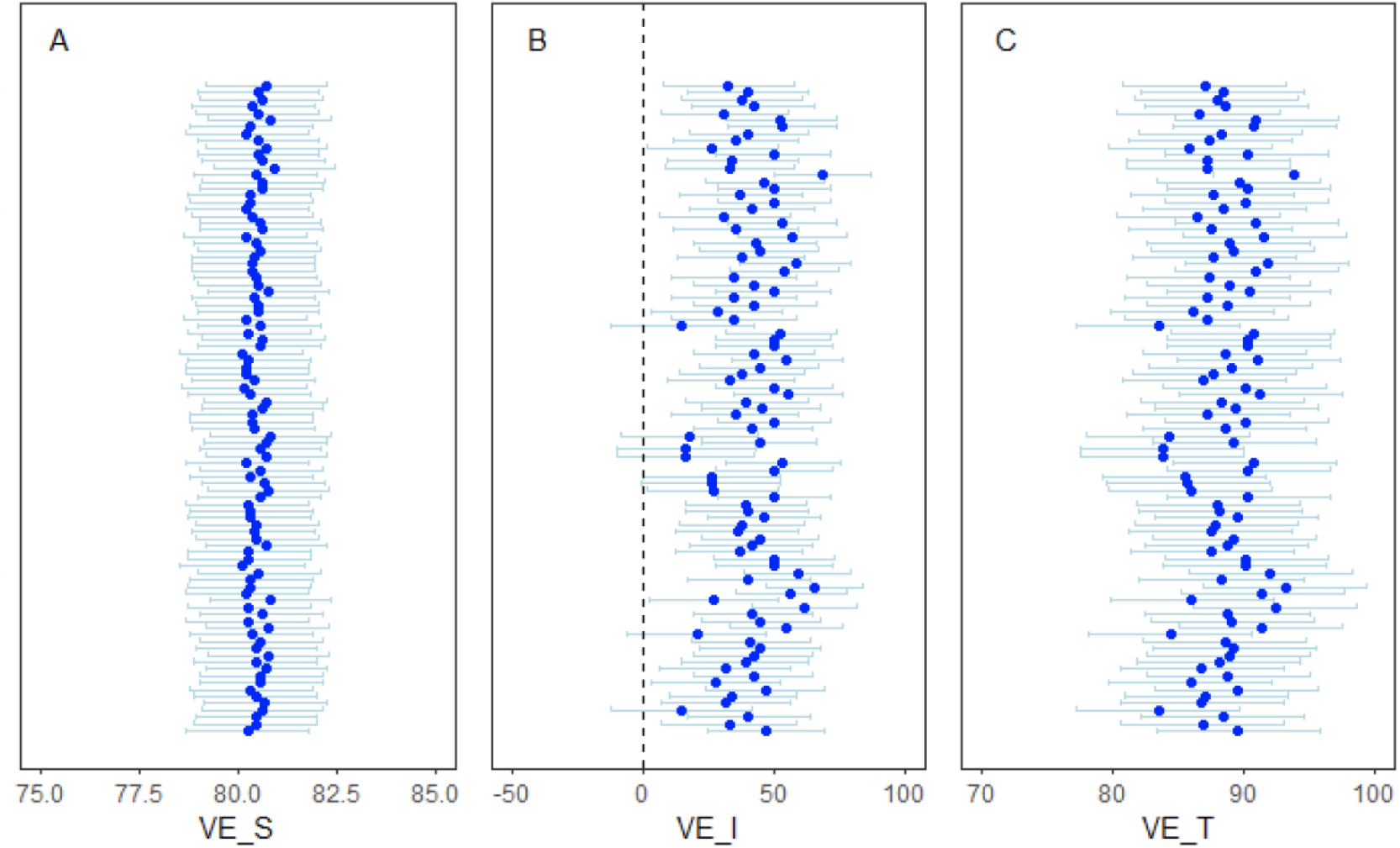
Forest-plot of the age-adjusted vaccine effectiveness estimates across the 100 iterations of the delay distributions from the primary transmission model. (**A**) Age-adjusted vaccine effectiveness against susceptibility to infection (***VE***_***S***_); (**B**) age-adjusted vaccine effectiveness against infectiousness given infection (***VE***_***I***_); (**C**) age-adjusted vaccine effectiveness against transmission (***VE***_***T***_).

**Fig. 3.**
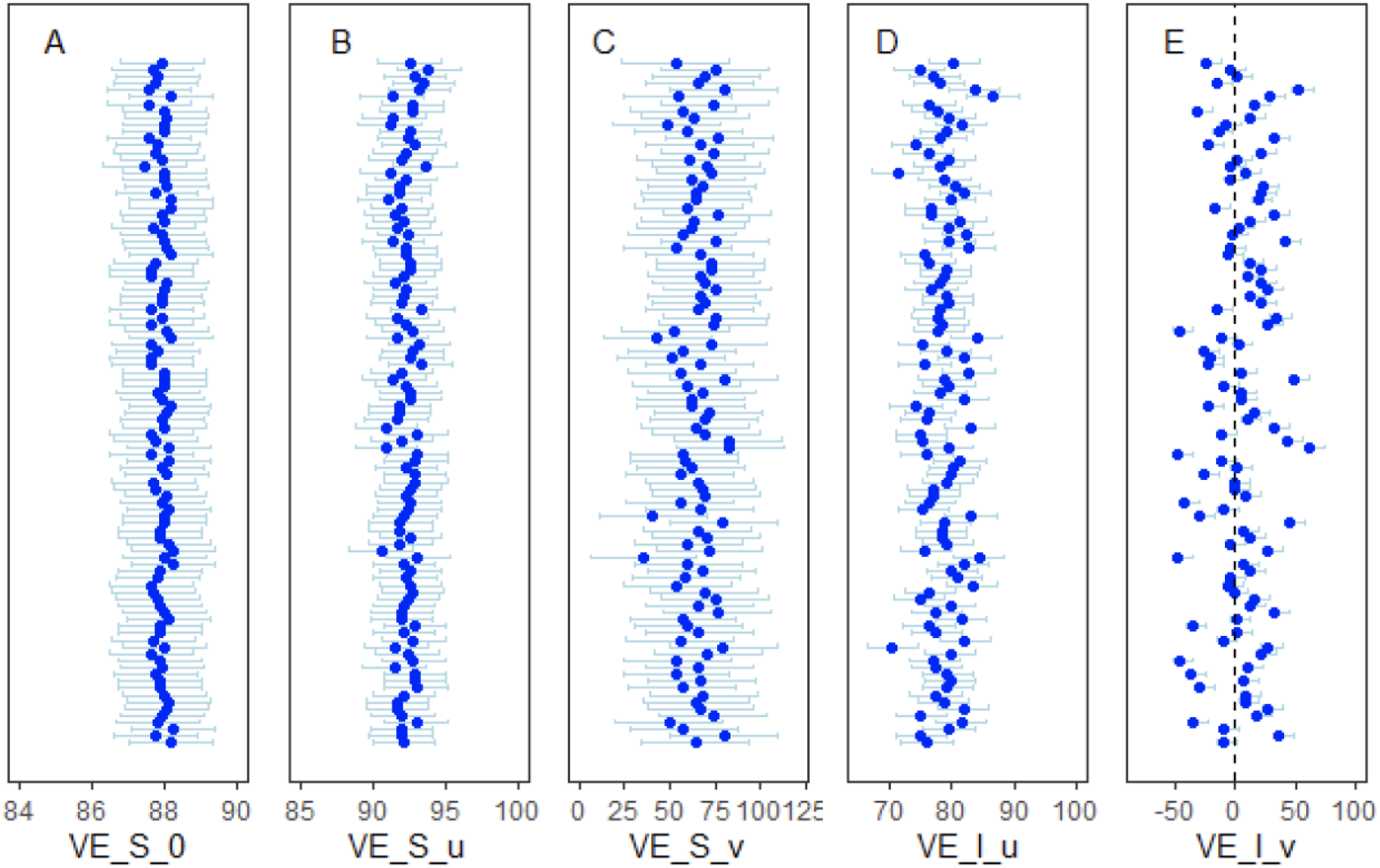
Forest-plot of the vaccine effectiveness estimates across the 100 iterations of the delay distributions from the alternative infection-hazard model. Vaccine effectiveness estimates against susceptibility to infection are plotted (**A**) in the absence of infected household members (***VE***_***S***,**0**_), or with at least one (**B**) unvaccinated (***VE***_***S***,***u***_) or (**C**) fully vaccinated household member (***VE***_***S***,***ν***_). The vaccine effectiveness estimates of being exposed to a fully vaccinated versus an unvaccinated infectious household member are plotted given (**D**) individual *j* is unvaccinated (*VE*_*I,u*_) or (**E**) individual *j* is fully vaccinated (*VE*_*I,v*_).

To date, there is limited published evidence with which to compare our estimates of vaccine effectiveness against infectiousness and transmission. A recent study of over 550,000 households in England showed vaccination with both the ChAdOx1 nCoV-19 and BNT162b2 vaccines reduced the odds of transmission from a vaccinated and infected household member by 40-50% compared to unvaccinated index cases [1, 4]. In previous studies, the index case in each household was defined as the earliest case of laboratory-confirmed COVID-19, by diagnosis date, and all secondary infections in the household were attributed to the index case [4]. In contrast, by inferring the date of infection, we do not assume that the index case in the household was necessarily the first individual to be diagnosed, and we account for the risk of transmission from other infected household members and from the community. Other studies investigating the reduction in infection risk among household members of vaccinated versus unvaccinated healthcare workers were conducted in Scotland and Finland, and provide indirect evidence of a lower risk of infection among household contacts of vaccinated individuals [1, 6, 13].

The two modeling approaches used in this study have different strengths and weaknesses. Both models adjusted for age, time-varying risk from the community, and the vaccination status of both the individual and other infected household members. However, the models differ in how they account for the contribution of other infected household members and the time-varying risk from the community. The vaccine effectiveness measures are naturally derived from the household transmission model, with a straightforward interpretation of the vaccine effectiveness against susceptibility to infection and against infectiousness given infection. The alternative model instead provides a case-by-case description of the reduction in risk depending on the vaccination status of the source(s) of exposure (for measures of the vaccine effectiveness against susceptibility to infection) and of the individual (for measures of vaccine effectiveness against infectiousness given the infection of other household members). Thus, the two models provide different perspectives of the reduction in risk following vaccination, which cannot be directly compared. Nevertheless, both approaches estimate a considerable reduction in both susceptibility to infection and infectiousness given infection following vaccination.

This study has several important limitations. Information on the true infection times (and duration of infectiousness) of infected household members is missing. To overcome this limitation, we sampled from three delay distributions parameterized from the literature to determine the potential infection status of each individual through time. Also, individuals who were infected but did not receive a SARS-CoV-2 test would be misclassified in our dataset. However, this is likely to have only a minor impact on our estimates (see supplementary material, table S7 and S8). We restricted our analysis to households with at least one infected individual and two or more household members (including those who never tested positive), which could bias estimates of the community force of infection [14]. However, since our primary goal was to determine the reduction in the relative risk of transmission following vaccination, and not to estimate the probability of transmission from the community versus infected household members, the decision to exclude households with no confirmed SARS-CoV-2 infections and/or fewer than two members is unlikely to bias our results. We nevertheless conducted a sensitivity analysis including 10,000 randomly select households with no infections and found that the results were robust to the inclusion of these households (table S9 and S10).

The ability of widespread vaccination to confer population-level protection through herd immunity depends on the vaccine effectiveness against transmission. Vaccination can prevent transmission by both providing protection against infection and reducing the infectiousness of vaccinated individuals who do become infected. Neither of these are typically directly measured in vaccine trials. By analyzing data on confirmed SARS-CoV-2 infections among household members in Israel, we provide measures of effectiveness of BNT162b2 against susceptibility to infection and against infectiousness given infection using two different approaches. Both models show evidence of a reduction in the infectiousness of vaccinated individuals who become infected in addition to protection against susceptibility to infection, leading to an overall reduction in the risk of transmission. Evidence of a high vaccine effectiveness against transmission confirms the importance of vaccinating both individuals at high and low risk of severe complications due to COVID-19 in order to maximize the population-level impact of vaccination and potentially achieve herd immunity.

## Data Availability

According to the IMOH (Israeli Ministry of Health) regulations, individual-level data cannot be shared openly. Specific requests for remote access to deidentified data should be referred to the Maccabi Institute for Research & Innovation.

## Author contributions

Each author’s contribution(s) to the paper should be listed [we encourage you to follow the <u>CRediT</u> model]. Each CRediT role should have its own line, and there should not be any punctuation in the initials.

Conceptualization: SG, TP, DMW, VEP

Methodology: OP, JLW, FWC, DMW, VEP

Investigation: OP, JLW, FWC, DMW, VEP

Visualization: OP, VEP

Funding acquisition: JLW, DMW, VEP

Project administration: SG, DMW, VEP

Supervision: DMW, VEP

Writing – original draft: OP

Writing – review & editing: OP, JLW, FWC, SG, TP, DMW, VEP

## Data and materials availability

According to the IMOH (Israeli Ministry of Health) regulations, individual level data cannot be shared openly. Specific requests for remote access to deidentified data should be referred to the Maccabi Institute for Research & Innovation.

## Materials and Methods

### Setting

Vaccination in Israel began on December 20, 2020, mainly using the BioNTech-Pfizer BNT162b2 vaccine, with a few individuals receiving the vaccine earlier. The vaccination campaign first targeted high-risk individuals, including those 60 years of age and older, medical personnel, workers at nursing homes, and individuals with comorbidities. After this phase, which lasted until January 21, age restrictions were lowered. By February 6, 2021, every Israeli citizen above 16 years old was eligible for the vaccine [10]. By the beginning of April 2021, 61% of the population had received at least one dose of the BNT162b2 vaccine [15].

The vaccination roll-out coincided with Israel’s third and largest wave of SARS-CoV-2 registered cases [16]. Consequently, a third national lockdown was issued in Israel starting December 24, 2020, with more severe restrictions (e.g., schools closures) issued starting January 8. These restrictions were progressively lifted starting on February 7, 2021.

### Data sources

We used data from Maccabi Healthcare Services (MHS) centralized computerized database, which captures all data on members’ healthcare-related interactions (including demographics, inpatient and outpatient visits, diagnoses, procedures etc). MHS is a nationwide 2.5 million-member state-mandated, not-for-profit sick fund in Israel, representing a quarter of the Israeli population, and is a representative sample of the Israeli population. The individual-level data for cases and household contacts include demographic information (i.e. age, sex), date of any polymerase chain reaction (PCR) tests for SARS-CoV-2 and the result of the test (considering that all such tests of MHS members are recorded centrally), and date of receipt of the first and second doses of the vaccine (if received). Individuals were defined as unvaccinated if they had not received any doses of BNT162b2 and fully vaccinated if at least 10 days had passed since receiving the second dose of the vaccine.

Due to computational constraints, we focused on households with at least one infected individual and two or more members; it was not possible to include all households with no infections. Sensitivity analyses were conducted using a randomly-selected subset of households with no infections to evaluate possible biases of this approach. We restricted our analysis to data from June 15, 2020 to March 24, 2021, since viral testing was not widely available prior to this date.

### Data augmentation for inferring transmission rates

For each individual, we observed the date at which the viral test was performed and the outcome of the test, but we do not have data on date of infection. To infer transmission rates, it is necessary to estimate when each individual within a household was infected and the period when each person was infectious. We therefore used a data augmentation approach to impute when a person with a positive PCR test was infected and infectious. This was accomplished using random samples from three different Gamma distributions representing the delay between onset of infectiousness and the date of the PCR test (i.e., *τ*_*report*_), between the date of infection and the onset of infectiousness (i.e., latent period, *τ*_*latent*_), and the time from the onset of infectiousness to the end of infectiousness (i.e., infectious period, *τ*_*infections*_). The values for these distributions were based on prior knowledge derived from observational studies on the latent period, times to seeking a test, and the duration of infectiousness (table S2) [17, 18]. For each individual, a random draw was taken from each of these distributions, and this process was repeated 100 times. For clarity, we refer to these Gamma distributions as the *delay* distributions.

For each person *j* in household *i* with a positive PCR test, let *T*_*ij*_ be the (imputed) time of infection in days since the beginning of the study: 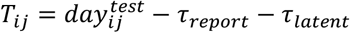, with 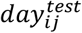 being the number of days after the start of the study until the PCR test date. We set the beginning of the study as May 29, 2020, since we allow for infections occurring up to 17 days prior to the start of the data (i.e., June 15, 2020). If no infection occurred for person *j, T*_*ij*_ is censored and equal to the total number of days in the study (i.e., *t*_*max*_). For the purposes of model fitting, we define *Y*_*ijt*_ as a binary variable equal to zero for each day of the study up until the time of infection (*t* = 1, …, *T*_*ij*_ − 1), equal to one for *T*_*ij*_ (assuming the person is infected), and censored from that point onwards (i.e., that person is relevant only in terms of transmitting to other household members). For people who are never infected, *Y*_*ijt*_ is equal to zero for all days of the study.

### Statistical modeling

Using the augmented data, we developed two discrete time-to-event data models of household transmission to estimate vaccine effectiveness against susceptibility to infection and against infectiousness given infection. In both approaches, we model the infection status for person *j* in household *i* on study day *t* (i.e., *Y*_*ijt*_) using conditionally independent Bernoulli distributions with corresponding probability of infection *π*_*ijt*_. These probabilities are then defined based on personal demographics, community risk, vaccination status, and characteristics of household transmission, with the approaches differing in how transmission is described. Both models were fit 100 times with different draws from the delay distributions to assess uncertainty in the results due to uncertainty in the unobserved dates of infection and infectiousness.

### Household transmission model

For the primary transmission model, we define the probability of infection on a given day as

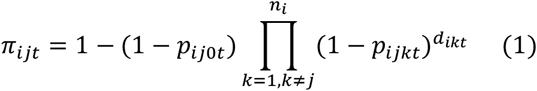

where *n*_*i*_ is the number of members in household *i, p*_*ij*0*t*_ is the probability that person *j* in household *i* is infected by the community on study day *t* (i.e., community risk of infection), *p*_*ijkt*_ is the probability that person *j* is infected by household member *k* (i.e., household risk of infection), and *d*_*ikt*_ is an indicator of whether person *k* can transmit to *j* on day *t*. In other words, 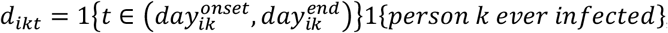, with 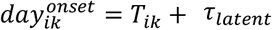 and 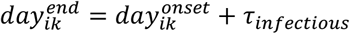 equal to the time of onset and end of infectiousness, respectively, for person *k*, and 1(.) representing the indicator function taking the value of one if the input condition is true and the value of zero otherwise.

The probability that individual *j* never tested positive for SARS-CoV-2 is given as

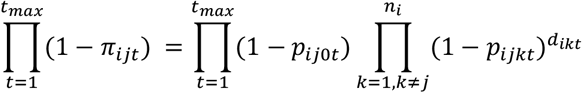

whereas the probability that individual *j* is infected on day *t*^*^ (before the end of the study) is given as

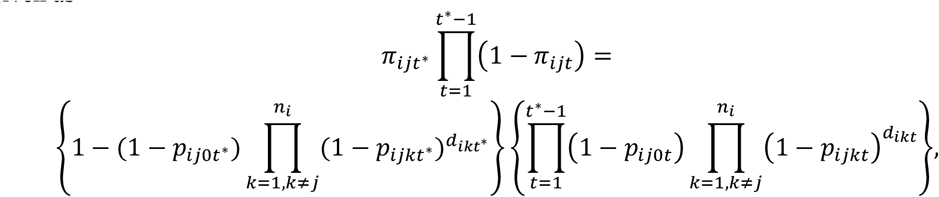

(i.e., the probability of escaping infection up to time (*t*^*^ − 1) multiplied by the probability of not escaping infection at time *t*^*^).

Conveniently (with respect to computation), the likelihood function can be written in terms of the introduced binary variables such as

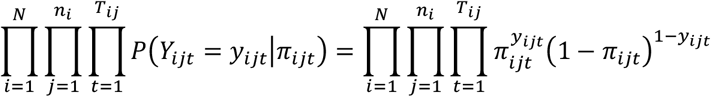

with *N* being the total number of households.

We define the per-person, per-day community risk of infection using the logit link function as

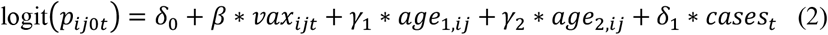

where *δ*_0_ is the baseline risk of infection from the community, *vax*_*ijt*_ is a binary variable equal to one if at least 10 days has passed since person *j* received the second dose of the vaccine, *age*_1,*ij*_ is a binary variable equal to one if the person is between 10 and 60 years old at the start of the study (reference category is the ≤10-year-old age group), *age*_2,*ij*_ is similarly defined for those aged ≥60 years old, and *cases*_*t*_ describes the time-varying risk from the community and is computed as the standardized number of positive PCR tests on day *t* in the data.

Similarly, the per-person, per-day risk of transmission from an infectious individual *k* to a susceptible household member *j* is defined (for *k*≠*j*) as

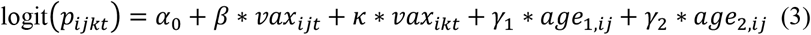

where *α*_0_ is the baseline risk of infection from an infected household member and *vax*_*ikt*_ is the vaccination status of household member *k*. All other terms have been previously described.

Vaccine effectiveness is expressed as a percentage and computed as 100*(1-*RR)*, with *RR* defined as a risk ratio comparing vaccinated and unvaccinated individuals respectively. Vaccine effectiveness against susceptibility stratified by age group was defined as

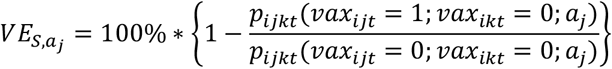

with *a*_*j*_ being a categorical variable equal to 0 for ages ≤10-year-old, 1 for ages between 10 and 60, and 2 for ages ≥60 years old; *vax*_*ijt*_ and *vax*_*ikt*_ are as previously defined. The explicit equation for age group 0 would then be

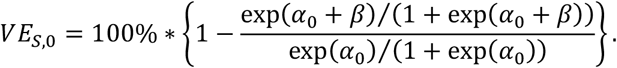

Including *γ*_1_ in both the numerators and denominators of the *VE*_*S*_ provides the VE with respect to age groups 1 (*VE*_*S*,1_) and similarly with *γ*_2_ for the VE with respect to age group 2 (*VE*_*S*,2_).

Vaccine effectiveness against infectiousness given infection is defined as

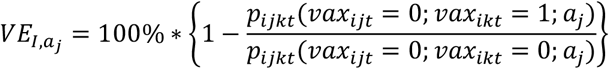

which is based on the vaccination status of infected household member *k*. Again, for age group 0, this is equivalent to

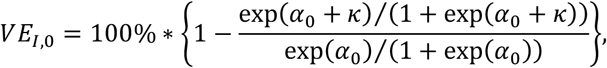

similarly, with *γ*_1_ and *γ*_2_ included for the other age groups.

We also estimated the vaccine effectiveness against transmission as

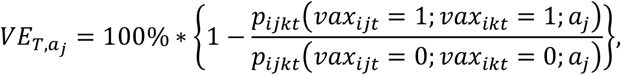

(i.e., the reduction in risk associated with vaccine-derived protection against both infection and infectiousness given infection). This is equivalent to

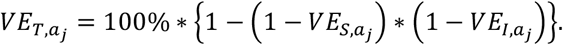

The age-adjusted vaccine effectiveness measures can be derived as 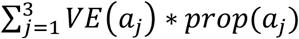, with *prop*(*a*_*j*_) being the fraction of people in each age group.

We initially applied the household transmission model to only households with a single occupant to inform the values of the baseline risk (*δ*_0_) and the time-varying risk from the community (*δ*_1_). We found *δ*_0_=-8.25 and *δ*_1_=0.79 and used these values as initial conditions for our analysis on households with at least one infected individual and two or more household members.

### Infection-hazard regression model

We compared the primary household transmission model defined by equations (1-3) with an alternative infection-hazard model. The models differ in their definition of the probability of infection, *π*_*ijt*_. We note that similar to the primary transmission model, the likelihood can be defined using the introduced binary variables, *Y*_*ijt*_. In this analysis, we use the complementary log-log-link function to connect the probabilities to individual-, household-, and community-level risk factors, such that

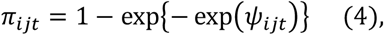

and

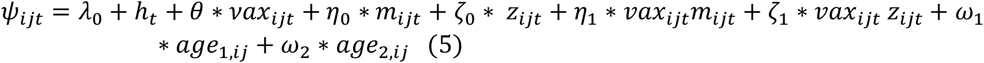

where *vax*_*ijt*_, *age*_1,*ij*_, and *age*_2,*ij*_ are as previously defined, *λ*_0_ is the intercept parameter, *h*_*t*_ is a smooth function of study time (modeled using splines) that describes the time-varying community risk, *m*_*ijt*_ is a binary variable equal to 1 if at least one other *unvaccinated* household member is infectious during study time *t* (not including person *j*), and *z*_*ijt*_ is a binary variable equal to 1 if at least one other *vaccinated* household member is infectious (not including person *j*). Interactions between person *j*’s vaccination status and the household risk variables are also included.

Use of the complementary log-log-link function leads to a hazard ratio interpretation for the exponentiated regression parameters. We define multiple susceptibility vaccine effects using this model output based on comparing different within-individual and within-household scenarios.

First, we compute

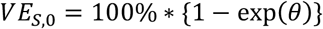

as the vaccine effectiveness against susceptibility to infection given there are no other infectious household members at that time (i.e., *m*_*ijt*_ = *z*_*ijt*_ = 0) and interpret it as the decrease in the hazard of infection due to vaccination. Similarly, we define

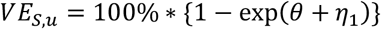

and

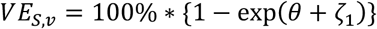

as the vaccine effectiveness against susceptibility to infection given there is at least one unvaccinated (*VE*_*S,u*_) or vaccinated (*VE*_*S,v*_) member in the household.

Vaccine effectiveness against infectiousness given the infection of household contacts is defined as

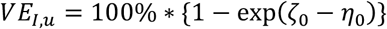

and

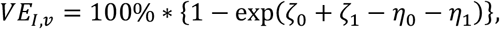

which can be interpreted as the percent reduction in the hazard of infection for individual *j* when exposed to a vaccinated versus unvaccinated infectious household member *k*, given individual *j* is unvaccinated (*VE*_*I,u*_) or vaccinated (*VE*_*I,v*_).

For both models, we summarized the vaccine effectiveness estimates by taking the mean over the 100 samples of the delay distributions. We derived the 95% confidence intervals (CI) using the law of total variance. All analyses were carried out in the R statistical software [19].

### Sensitivity analysis: including the first vaccine dose

For the primary household transmission model, we included information on the first vaccine dose status by defining community risk as

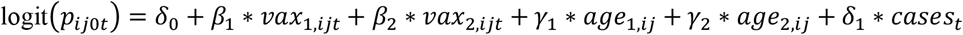

where *vax*_1,*ijt*_, and *vax*_2,*ijt*_represent mutually exclusive binary variables of vaccination status (i.e., individual *j* in household *i* is partially or fully vaccinated at time *t*, respectively), and all other terms have been previously described.

Similarly, household risk is defined (for *k*≠*j*) as:

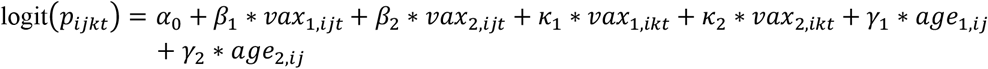

where *vax*_1,*ikt*_ is the first-dose vaccination status of household member *k* at time *t* and *vax*_2,*ikt*_ is the second-dose vaccination status of household member *k* at time *t*. Vaccine effectiveness estimates are the same as for the primary analysis.

For the infection-hazard model, we now define the variable *ψ*_*ijt*_ from equations 4-5 as

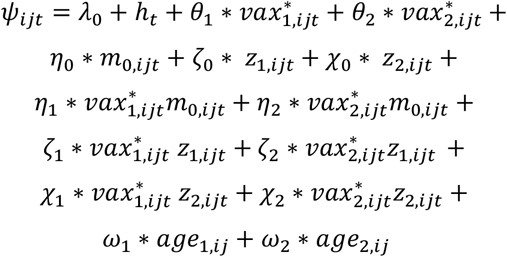

where 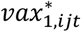, and 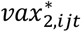 represent mutually exclusive binary variables of vaccination level (i.e., partially and fully vaccinated, respectively), *m*_0,*ijt*_ is a binary variable equal to one if at least one other *unvaccinated* household member is infectious during study time *t* (not including person *j*), *z*_1,*ijt*_ is a binary variable equal to one if at least one other *partially vaccinated* household member is infectious (not including person *j*), and *z*_2,*ijt*_ is a binary variable equal to one if at least one other *fully vaccinated* household member is infectious (not including person *j*). Interactions between person *j*’s vaccination status and household risk are also included.

The vaccine effectiveness against susceptibility to infection given there are no other infectious household members at that time (i.e., *m*_0,*ijt*_ = *z*_1,*ijt*_ = *z*_2,*ijt*_ = 0) is given as:

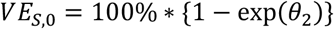

and is interpreted as the decrease in the hazard of infection due to full vaccination.

Similarly, we define

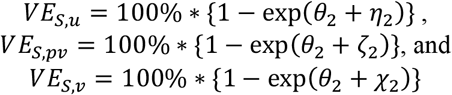

as the vaccine effectiveness against susceptibility to infection given there is at least one unvaccinated (*VE*_*S,u*_), partially vaccinated (*VE*_*S,pv*_), or fully vaccinated (*VE*_*S,v*_) member in the household.

Vaccine effectiveness against infectiousness given the infection of household contacts is defined as

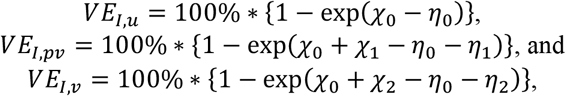

which can be interpreted as the effect of being exposed to a fully vaccinated versus unvaccinated infectious household member given individual *j* is unvaccinated (*VE*_*I,u*_), partially vaccinated (*VE*_*I,pv*_), or fully vaccinated (*VE*_*I,v*_). Results from both models are reported in tables S4 and S5.

### Sensitivity analysis: including a subset of households with no infections

We randomly selected 10,000 households with at least two household members and no detected infections and included them along with our original set of households with at least two household members and at least one infection. We compared the vaccine effectiveness results from both models for one iteration of the delay distributions (tables S6 and S7).

### Sensitivity analysis: testing the robustness of the results to misclassification of cases

We run a sensitivity analysis to test the robustness of the results to misclassification of individuals who were infected but did not receive a SARS-CoV-2 test. For each individual with a negative PCR test, we randomly selected a new PCR test date. On this new PCR test date, each individual could now have a positive PCR test based on a Bernoulli distribution with probability *p* = *α* * *prob*_*test*_. We define *prob*_*test*_ = (total number of positive PCR cases in the dataset)/(total population in the dataset) = 0.08. With this new dataset, we ran the delay distribution process for one iteration, and we estimated the vaccine effectiveness from both models. We tested two scenarios: (a) *α* = 0.01 and (b) *α* = 0.10 (tables S8 and S9).

**Table S1.** Distribution of PCR-confirmed SARS-CoV-2 infections across households of varying size.

| **NUMBER OF**<br>**INFECTED HOUSEHOLD**<br>**MEMBERS** | **<u>NUMBER OF HOUSEHOLD MEMBERS</u>** |  |  |  |  | **TOTAL (%)** |
| --- | --- | --- | --- | --- | --- | --- |
|  | **1** | **2** | **3** | **4** | **5+** |  |
| **0** | 714451 | 211419 | 90539 | 72872 | 60558 | 1149839 (90.2) |
| **1** | 59552 | 11874 | 7299 | 6039 | 7666 | 92430 (7.2) |
| **2** |  | 7737 | 3440 | 2702 | 3180 | 17059 (1.3) |
| **3** |  |  | 2809 | 1994 | 2336 | 7139 (0.6) |
| **4** |  |  |  | 1870 | 2154 | 4024 (0.3) |
| **5+** |  |  |  |  | 4524 | 4524 (0.4) |
| **TOTAL (%)** | 774003 (60.7) | 231030 (18.1) | 104087 (8.2) | 85477 (6.7) | 80418 (6.3) | 1275015 |

**Table S2.** Delay distribution parameters for the data augmentation process.

| **Parameter** | **Description** | **Value** | **Reference** |
| --- | --- | --- | --- |
| $\tau_{report}$ | distribution for the time from onset of infectiousness to testing | $\sim \text{Gamma}(\text{shape}=2.34; \text{scale}=1/2.59)$ | [18] |
| $\tau_{infectious}$ | distribution for the infectious period | $\sim \text{Gamma}(\text{shape}=4; \text{scale}=5/4)$ | [17] |
| $\tau_{latent}$ | distribution for the latent period | $\sim \text{Gamma}(\text{shape}=4; \text{scale}=1)$ | [17] |

**Table S3.** Vaccine effectiveness estimates by age group from the primary transmission model.

| Age group | Type of vaccine effectiveness measure | Estimate [95% confidence interval] |
| --- | --- | --- |
| *Vaccine effectiveness against susceptibility to infection* |  |  |
| 0-10y | $VE_{S,1}$ | 80.6% [79.0%, 82.3%] |
| 11-59y | $VE_{S,2}$ | 80.4% [78.7%, 82.1%] |
| 60y+ | $VE_{S,3}$ | 80.2% [78.5%, 81.8%] |
| Overall | $VE_{S,adjusted}$ | 80.5% [78.9%, 82.1%] |
| *Vaccine effectiveness against infectiousness given infection* |  |  |
| 0-10y | $VE_{I,1}$ | 41.5% [9.1%, 73.9%] |
| 11-59y | $VE_{I,2}$ | 41.2% [8.9%, 73.5%] |
| 60y+ | $VE_{I,3}$ | 41.0% [8.6%, 73.1%] |
| Overall | $VE_{I,adjusted}$ | 41.3% [9.5%, 73.0%] |
| *Vaccine effectiveness against transmission* |  |  |
| 0-10y | $VE_{T,1}$ | 88.7% [82.4%, 95.0%] |
| 11-59y | $VE_{T,2}$ | 88.5% [82.1%, 94.8%] |
| 60y+ | $VE_{T,3}$ | 88.3% [82.0%, 95.0%] |
| Overall | $VE_{T,adjusted}$ | 88.5% [82.3%, 94.8%] |

**Table S4.** Description of the parameter estimates on the odds ratio (OR) or inverse logit (IL) scale with 95% confidence intervals (CI) averaged over the 100 iterations of the delay distributions from the primary transmission model.

| Parameter | Description | OR/IL [95%CI] |
| --- | --- | --- |
| $\frac{\exp(\alpha_0)}{1 + \exp(\alpha_0)}$ | baseline probability of transmission per day from an infected HH member to a susceptible child (i.e., ref category) | 0.016 [0.015, 0.016] |
| $\frac{\exp(\delta_0)}{1 + \exp(\delta_0)}$ | baseline probability of transmission per day from the community to a susceptible child (i.e., ref category) | 0.0010 [0.0009, 0.0010] |
| $\exp(\beta)$ | vaccination status of the individual | 0.19 [0.18, 0.21] |
| $\exp(\kappa)$ | vaccination status of other HH members | 0.57 [0.38, 0.88] |
| $\exp(\gamma_1)$ | age >10 and <60 | 1.83 [1.80, 1.85] |
| $\exp(\gamma_2)$ | ages $\geq 60$ | 2.77 [2.71, 2.83] |
| $\exp(\delta_1)$ | time-varying risk from the community | 1.77 [1.77, 1.78] |

**Table S5.** Vaccine effectiveness estimates from the primary transmission model across age groups including both vaccine doses.

| Age group | Type of vaccine effectiveness measure | Estimate [95% confidence interval] |
| --- | --- | --- |
| *Vaccine effectiveness against susceptibility to infection* |  |  |
| 0-10y | $VE_{S,1}$ | 80.9% [79.3%, 82.5%] |
| 11-59y | $VE_{S,2}$ | 80.7% [79.1%, 82.3%] |
| 60y+ | $VE_{S,3}$ | 80.5% [78.9%, 82.1%] |
| Overall | $VE_{S,adjusted}$ | 80.8% [79.6%, 82.0%] |
| *Vaccine effectiveness against infectiousness given infection* |  |  |
| 0-10y | $VE_{I,1}$ | 42.6% [18.7%, 66.4%] |
| 11-59y | $VE_{I,2}$ | 42.2% [18.4%, 66.0%] |
| 60y+ | $VE_{I,3}$ | 41.9% [18.1%, 65.6%] |
| Overall | $VE_{I,adjusted}$ | 42.3% [19.0%, 65.6%] |
| *Vaccine effectiveness against transmission* |  |  |
| 0-10y | $VE_{T,1}$ | 89.0% [84.4%, 93.7%] |
| 11-59y | $VE_{T,2}$ | 88.9% [84.2%, 93.5%] |
| 60y+ | $VE_{T,3}$ | 88.6% [83.9;93.4] |
| Overall | $VE_{T,adjusted}$ | 88.9% [84.4;93.5] |

**Table S6.** Vaccine effectiveness estimates from the alternative infection-hazard model including both vaccine doses.

| Interpretation | Type of VE | Estimate [95% confidence interval] |
| --- | --- | --- |
| *Vaccine effectiveness against susceptibility to infection* |  |  |
| in the absence of infected household members | $VE_{S,0}$ | 89.3% [88.3%, 90.3%] |
| with at least one unvaccinated household member | $VE_{S,u}$ | 92.3% [90.5%, 94.0%] |
| with at least one partially vaccinated household member | $VE_{S,pv}$ | 91.6% [84.1%, 99.1%] |
| with at least one fully vaccinated household member | $VE_{S,v}$ | 75.8% [47.5%, 104.1%] |
| *Effect of being exposed to a fully vaccinated vs unvaccinated infectious household member* |  |  |
| given individual $j$ is unvaccinated | $VE_{I,u}$ | 89.8% [86.4%, 93.1%] |
| given individual $j$ is partially vaccinated | $VE_{I,pv}$ | 85.5% [71.0%, 100%] |
| given individual $j$ is vaccinated | $VE_{I,v}$ | 68.0% [31.7%, 104.3%] |

**Table S7.** Vaccine effectiveness estimates from the primary transmission model across age groups testing for misclassified cases with scenario a) *α* = 0. 01; scenario b) *α* = 0. 10.

| Age group | Type of vaccine effectiveness measure | a) Estimate [95% confidence interval] | b) Estimate [95% confidence interval] |
| --- | --- | --- | --- |
| *Vaccine effectiveness against susceptibility to infection* |  |  |  |
| 0-10y | $VE_{S,1}$ | 80.5% [78.9%, 82.1%] | 79.6% [78.0%, 81.3%] |
| 11-59y | $VE_{S,2}$ | 80.3% [78.7%, 81.9%] | 79.4% [77.8%, 81.1%] |
| 60y+ | $VE_{S,3}$ | 80.1% [78.5%, 81.7%] | 79.2% [77.6%, 80.9%] |
| Overall | $VE_{S,adjusted}$ | 80.4% [78.8%, 81.9%] | 79.5% [77.9%, 81.1%] |
| *Vaccine effectiveness against infectiousness given infection* |  |  |  |
| 0-10y | $VE_{I,1}$ | 32.5% [7.1%, 58.0%] | 41.6% [18.9%, 64.3%] |
| 11-59y | $VE_{I,2}$ | 32.2% [6.9%, 57.6%] | 41.3% [18.6%, 63.9%] |
| 60y+ | $VE_{I,3}$ | 31.9% [6.7%, 57.1%] | 40.9% [18.4%, 63.5%] |
| Overall | $VE_{I,adjusted}$ | 32.3% [7.8%, 56.8%] | 41.4% [19.5%, 63.3%] |
| *Vaccine effectiveness against transmission* |  |  |  |
| 0-10y | $VE_{T,1}$ | 86.9% [81.8%, 92.0%] | 88.1% [83.4%, 92.9%] |
| 11-59y | $VE_{T,2}$ | 86.7% [81.5%, 91.8%] | 87.9% [83.2%, 92.7%] |
| 60y+ | $VE_{T,3}$ | 86.4% [81.3%, 91.6%] | 87.7% [82.9%, 92.5%] |
| Overall | $VE_{T,adjusted}$ | 86.7% [81.8%, 91.7%] | 88.0% [83.3%, 92.6%] |

**Table S8.** Vaccine effectiveness estimates from the alternative hazard model testing for misclassified cases with scenario a) *α* = 0. 01; scenario b) *α* = 0. 10.

| Interpretation | Type of VE | a) Estimate [95% confidence interval] | b) Estimate [95% confidence interval] |
| --- | --- | --- | --- |
| ***Vaccine effectiveness against susceptibility to infection*** |  |  |  |
| in the absence of infected household members | $VE_{S,0}$ | 87.7% [86.6%, 88.8%] | 87.2% [86.1%, 88.3%] |
| with at least one unvaccinated household member | $VE_{S,u}$ | 92.5% [90.7%, 94.3%] | 92.6% [90.9%, 94.3%] |
| with at least one fully vaccinated household member | $VE_{S,v}$ | 61.3% [34.9%, 87.8%] | 66.9% [42.3%, 91.5%] |
| ***Effect of being exposed to a fully vaccinated vs unvaccinated infectious household member*** |  |  |  |
| given individual $j$ is unvaccinated | $VE_{I,u}$ | 82.2% [76.2%, 88.1%] | 82.2% [76.2%, 88.3%] |
| given individual $j$ is vaccinated | $VE_{I,v}$ | 8.2% [-53.4%, 69.7%] | 20.6% [-36.8%, 78.1%] |

**Table S9.**
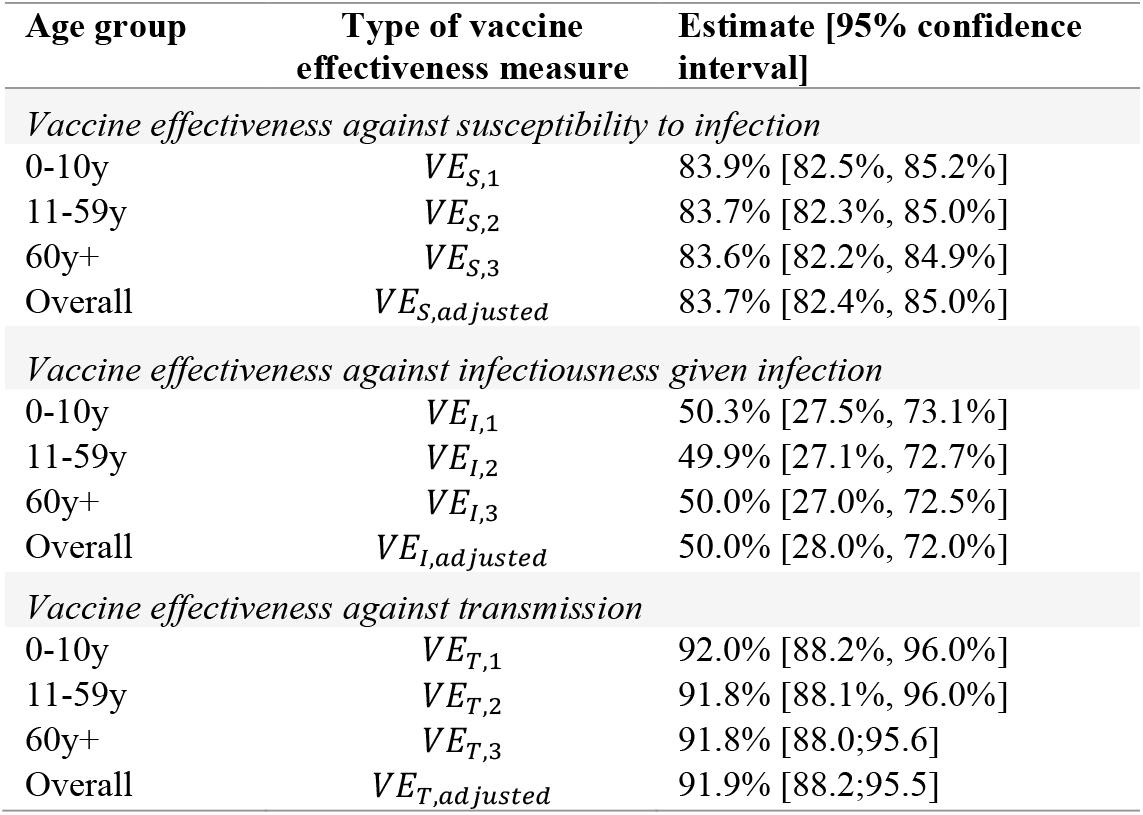
Vaccine effectiveness estimates from the primary transmission model across age groups including a subset of households with no infections.

| Age group | Type of vaccine effectiveness measure | Estimate [95% confidence interval] |
| --- | --- | --- |
| *Vaccine effectiveness against susceptibility to infection* |  |  |
| 0-10y | $VE_{S,1}$ | 83.9% [82.5%, 85.2%] |
| 11-59y | $VE_{S,2}$ | 83.7% [82.3%, 85.0%] |
| 60y+ | $VE_{S,3}$ | 83.6% [82.2%, 84.9%] |
| Overall | $VE_{S,adjusted}$ | 83.7% [82.4%, 85.0%] |
| *Vaccine effectiveness against infectiousness given infection* |  |  |
| 0-10y | $VE_{I,1}$ | 50.3% [27.5%, 73.1%] |
| 11-59y | $VE_{I,2}$ | 49.9% [27.1%, 72.7%] |
| 60y+ | $VE_{I,3}$ | 50.0% [27.0%, 72.5%] |
| Overall | $VE_{I,adjusted}$ | 50.0% [28.0%, 72.0%] |
| *Vaccine effectiveness against transmission* |  |  |
| 0-10y | $VE_{T,1}$ | 92.0% [88.2%, 96.0%] |
| 11-59y | $VE_{T,2}$ | 91.8% [88.1%, 96.0%] |
| 60y+ | $VE_{T,3}$ | 91.8% [88.0;95.6] |
| Overall | $VE_{T,adjusted}$ | 91.9% [88.2;95.5] |

**Table S10.**
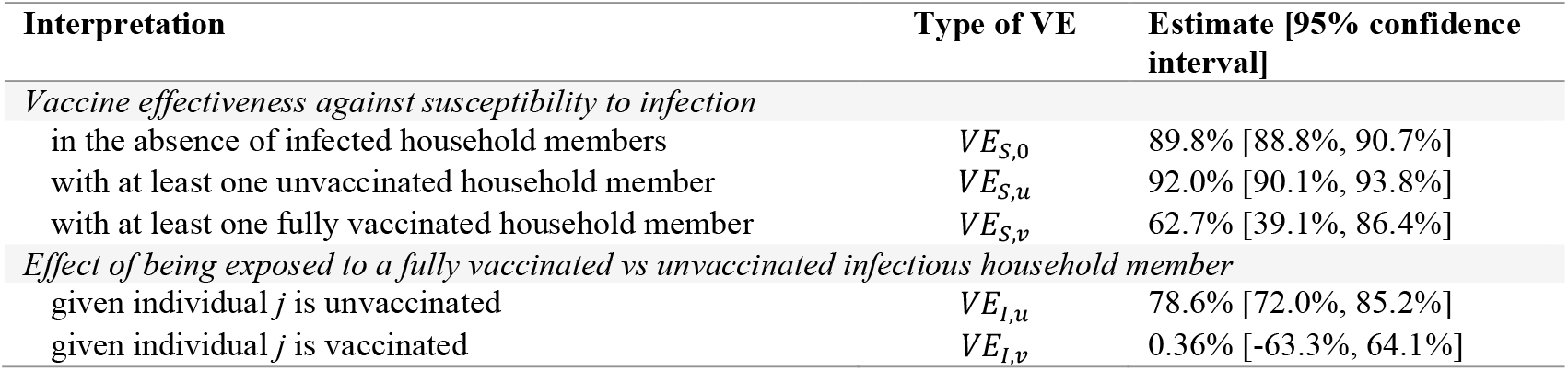
Vaccine effectiveness estimates from the alternative hazard model including a subset of households with no infections.

| Interpretation | Type of VE | Estimate [95% confidence interval] |
| --- | --- | --- |
| *Vaccine effectiveness against susceptibility to infection* |  |  |
| in the absence of infected household members | $VE_{S,0}$ | 89.8% [88.8%, 90.7%] |
| with at least one unvaccinated household member | $VE_{S,u}$ | 92.0% [90.1%, 93.8%] |
| with at least one fully vaccinated household member | $VE_{S,v}$ | 62.7% [39.1%, 86.4%] |
| *Effect of being exposed to a fully vaccinated vs unvaccinated infectious household member* |  |  |
| given individual $j$ is unvaccinated | $VE_{I,u}$ | 78.6% [72.0%, 85.2%] |
| given individual $j$ is vaccinated | $VE_{I,v}$ | 0.36% [-63.3%, 64.1%] |

